# Responsiveness of the electrically stimulated cochlear nerve in patients with a missense variant in *ACTG1*: Preliminary Results

**DOI:** 10.1101/2022.10.24.22281451

**Authors:** Yi Yuan, Denise Yan, Jeffrey Skidmore, Prem Chapagain, Xuezhong Liu, Shuman He

## Abstract

**Objectives:** This preliminary study identified a missense variant in *ACTG1* (NM_001614.5) in a family with autosomal dominant non-syndromic hearing loss (ADNSHL). The responsiveness of the electrically-stimulated cochlear nerve (CN) in two implanted participants with this missense change was also evaluated and reported.

**Design:** Genetic testing was done using a custom capture panel (MiamiOtoGenes) and whole exome sequencing. The responsiveness of the electrically-stimulated CN was evaluated in two members of this family (G1 and G4) using the electrically evoked compound action potential (eCAP). eCAP results from these two participants were compared with those measured three implanted patient populations: children with cochlear nerve deficiency, children with idiopathic hearing loss and normal-sized cochlear nerves, and postligually deafened adults.

**Results:** Sequencing of *ACTG1* identified a missense c.737A>T (p. Gln246Leu) variant in *ACTG1* (NM_001614.5) which is most likely the genetic cause of ADNSHL in this family. eCAP results measured in these two participants showed substantial variations.

**Conclusion:** The missense c.737A>T (p. Gln246Leu) variant in *ACTG1* (NM_001614.5) co-segregated with hearing loss in this family. The responsiveness of the electrically-stimulated CN can vary among patients with the same genetic variants, which suggests the importance of evaluating the functional status of the CN for individual CI patients.

## 1. Introduction

Autosomal dominant non-syndromic hearing loss (ADNSHL) accounts for approximately 20% of non-syndromic hereditary hearing loss cases, and is often characterized by a post-lingual onset and progressive hearing loss (Shearer et al., 1993). It has a broad phenotypic spectrum which includes considerable variation in age of onset, type of hearing loss configurations (i.e., low-frequency, mid-frequency, high-frequency, and all-frequency), and the rate of progression (see review by Pennings et al., 2003). Today, around 51 genes and 68 chromosomal loci have been associated with ADNSHL (Hereditary Hearing Loss Homepage; http://hereditaryhearingloss.org, 2021). These deafness genes encode a myriad of proteins that have a specialized role in various cell types, structures, and processes in the cochlea (Hilgert et al., 2009).

Much of the function of the inner ear relies on the hair bundle, a cluster of actin-filled stereocilia for transduction of auditory and vestibular stimuli into neural impulses. Mutations that alter the amino acid structure of actin are generally not tolerated, because actin has a large repertoire of interacting ligands and actin-binding partners to perform various functions, including movement, cellular structures, redistribution of surface receptors for regulation of cellular responses and other processes (e.g., Kovar et al., 2006; Ferron et al., 2007). Specifically, depending on the organization of the filaments, actin can be deployed for support of a variety of cellular processes. Microfilament functions include cell motility or control of cytokinesis (Pollard et al., 2009); cells also use actin to adopt and maintain specific cellular structures (e.g., Michelot et al., 2011). In the apical region of the hair cell, parallel bundles of actin fibers with identical polarity mediate protrusion of stereocilia (Tilney et al., 1983); in the cuticular plate lying beneath the apical surface, a dense meshwork of actin filaments anchors the stereocilia (DeRosier & Tilney, 1989) and the adherents junction encircling the hair cell next to the apical region also contains antiparallel actin filaments (Hirokawa &Tilney, 1982). Actin structures that support the plasma membrane of cells and provide a track for the passage of synaptic vesicles are also found in the basolateral region of the hair cell.

There are six actin genes in the human genome. Four of these encode the α-actin isoforms present in various muscle cells and the remaining two actin genes, of which one encoding the γ-actin isoform predominantly expressed (*ACTG1*) in the auditory hair cell, and β-actin (ACTB), an isoform at the front of the cell where actin filaments polymerize. The β- and γ-actin isoforms are present in non-muscle cells and differ at only four amino acids positions at the N-terminus. These actin isoforms are highly conserved at the protein level, with >90% homology between the single yeast actin (ACT1) gene and the human γ-actin. Vertebrate γ-actin amino acids are identical between humans, mice, cattle, and chickens (Sheterline et al., 1998).

*ACTG1* mutations were identified as responsible for either non-syndromic deafness or Baraitser-Winter syndrome, a disorder characterized by distinct craniofacial features, ocular colobomata and neuronal migration defect and variable hearing loss (e.g., Riviere et al., 2012). It is believed that mutations in γ-actin cause hearing loss mainly by impairing the function and/or viability of hair cells (Rendtorff et al., 2006; Van Wijk et al., 2003). DFNA20 (MIM 604717) and DFNA26 (MIM 604717) are loci linked to dominant, non-syndromic, progressive hearing loss that map to chromosome 17q25.3 (Morell et al., 2000; Yang & Smith, 2000). Individuals with the DFNA20 and DFNA26 (DFNA20/26) exhibit sensorineural hearing loss that, very similar to age related hearing loss, affects initially only the high frequencies and steadily progresses to include all frequencies (Yang & Smith, 2000; Elfenbein et al., 2001). Distortion product otoacoustic emission (DPOAE) assessments are consistent with a cochlear site of lesion (Elfenbein et al., 2001). However, Teig (1968) reported pathological tone decay at 2 and 4 kHz when hearing loss is more than 50 dB in patients with *ACTG1* missense mutations. These results suggested potential neural lesions along the auditory pathway in patients with *ACTG1* missense mutations.

Theoretically, neural encoding and processing of electrical stimulation in implanted patients who carry the *ACTG1* missense mutation should not be compromised since this genetic lesion mainly affects the hair cells which are “bypassed” by the cochlear implant (CI). As a result, patients with *ACTG1* missense mutation are expected to have good CI clinical outcomes. However, this theoretical possibility has not been evaluated in human CI users, which represents a knowledge gap in the field of cochlear implantation. The responsiveness of the electrically-stimulated CN has been suggested to be important for hearing outcomes in CI users (e.g., Long et al., 2014; He et al., 2018; Skidmore et al., 2021). Therefore, it was selected as the functional, phenotypic readout in this preliminary study.

In this study, we ascertained a five-generation family with progressive ADNSHL. We used our targeted custom MiamiOtoGenes panel containing 230 hearing loss-associated genes to analyze DNA samples from 2 affected family members (G2 and G4) and sample from 1 patient (G3) was subjected to whole exome sequencing (WES). Using this strategy, we identified a missense c.737A>T (p. Gln246Leu) variant in *ACTG1* (NM_001614.5) that co-segregated with hearing loss and most likely to be the genetic cause of ADNSHL in this family. This genetic finding provided an opportunity to explore the phenotypic effect of this missense change on the responsiveness of the CN to electrical stimulation. The responsiveness of the electrically stimulated CN was also evaluated using electrophysiological measures of the electrically evoked compound action potential (eCAP) in this preliminary study. The eCAP is a near-field measure of synchronized neural responses generated by a group of CN fibers. eCAP measures that can be used to assess the responsiveness of the CN to electrical stimulation include slope of the eCAP amplitude growth function (AGF), the absolute refractory period (*t*_0_), and the speed of recovery from neural adaption (*τ*_3_). The eCAP AGF represents a series of eCAP amplitudes recorded at corresponding stimulation levels (Skidmore et al., 2021). The rate at which the eCAP amplitude increases with increasing stimulation level (i.e., slope) has been shown to be associated with the responsiveness of CN fibers, with greater slopes indicating better CN responsiveness (He et al., 2018, 2020). *t*_0_ is an estimate of the absolute recovery period of the CN fibers based on the eCAP refractory recovery function (RRF). It represents the minimum time interval that is needed to elicit the eCAP to a second pulse (i.e., probe pulse) after a suprathreshold masking pulse (He et al., 2018; Skidmore et al., 2021). Smaller *t*_0_ values indicate better neural responsiveness in both animal models (Shepherd et al., 2004) and human CI users (He et al., 2018). *τ*_3_ has been shown to be associated with spiral ganglion neuron (SGN) survival in animal models (Ramekers et al., 2015). In human CI users, patients with longer *τ*_3_s tend to have poorer speech perception performance (He et al., 2022).

In this preliminary and descriptive study, we reported the identification of the missense c.737A>T (p. Gln246Leu) variant in *ACTG1* (NM_001614.5), and the responsiveness of the electrically stimulated CN evaluated in two participants with this missense change. In addition, their electrophysiological results were compared with three implanted patient populations: children with cochlear nerve deficiency (CND), children with idiopathic sensorineural hearing loss and normal-sized cochlear nerves (NSCNs) and postlingually deafened adult CI users with various etiologies of hearing loss, including trauma, noise exposure, Meniere’s disease, autoimmune disease, vestibular schwannoma, sudden hearing loss, etc.

## 2. Materials and Methods

### 2.1 Participants

We ascertained a five-generation American family with ADSNHL. Saliva samples were collected, and DNA was extracted from six available family members, of whom five were affected. All clinical data of affected members in these families were obtained from questionnaires and included age at onset, degree, evolution, and symmetry of the hearing loss, presence of tinnitus, and other relevant clinical manifestations. None of the patients had any history of chronic noise exposure or ototoxic drugs. The hearing levels of the participants were evaluated by pure-tone audiometry. Hearing information of this family is listed in Table 1.

**Table 1.** Hearing loss information of genetic testing participants. HL: hearing loss, SN: Sensorineural

| Subject Number | Hearing Status | Affected side | Type of HL | Progression of HL | Severity of HL | Audiogram |
| --- | --- | --- | --- | --- | --- | --- |
| G1 | Hearing impaired | Bilateral, symmetric | SN | Progressive | Profound (>95dB) | Flat (<15 dB diff. .25-.5, 1-2, 4-8 kHz) |
| G2 | Hearing impaired | Bilateral, symmetric | SN | Progressive | Mild (20-40dB) | High frequency gently sloping (15-29 dB diff. .5-1 and 4-8 kHz) |
| G3 | Hearing Impaired | NA | NA | NA | NA | NA |
| G4 | Hearing impaired | Bilateral, symmetric | SN | Progressive | Moderately severe - profound | Moderately severe sloping to profound (5-20 dB diff. .25-.5 and 1-2, 4-8 kHz) |
| G5 | Hearing impaired | Unilateral: R | SN | Sudden, Stable | Profound (>95dB) | Flat (<15 dB diff. .25-.5, 1-2, 4-8 kHz) |
| G6 | Normal hearing | NA | NA | NA | NA | NA |

Among the five affected participants, only G1 and G4 are CI users. As a result, electrophysiological evaluations were only conducted in these two participants. Both participants were implanted with a Cochlear™ Nucleus® (Cochlear Ltd., Macquarie, NSW, Australia) CI (24RE[CA]) with a full electrode array insertion in the test ear. The Cochlear™ Nucleus® CI has 22 CI electrodes, with electrode 1 typically located at the base and electrode 22 located toward the apex of the cochlea. G1 was implanted in both ears and G4 was unilaterally implanted in the right ear. All three implanted ears were assessed in this study. Electrode locations tested in each participant for each eCAP measure are listed in Table 2.

**Table 2.** Electrophysiological testing information of study participants. L: left, R: right, AGF: amplitude growth function, RRF: refractory recovery function, AR: adaptation recovery. *Electrode 12 was tested due to artifact contaminations of neural responses recorded at electrode 9.

| Subject number | Ear tested | Electrodes tested |  |
| --- | --- | --- | --- |
|  |  | AGF and RRF | AR |
| G1 | R | 3, 12 and 21 | 3,9,15 and 21 |
| G1 | L | 3, 12 and 21 | 3,9,15 and 21 |
| G4 | R | 3, 12 and 21 | 3,12*,15 and 21 |

The study was approved by the ethics committee of The Ohio State University (2017H0131 and 2018H0587). Written informed consent was obtained from all participants prior to saliva sample collection and electrophysiological evaluation.

### 2.2. Genetic Testing Procedures

#### 2.2.1. Targeted Genomic Capture and Whole Exome Sequencing

Genomic DNA was extracted from saliva samples using the commercial kit Oragene™ (DNA Genotek OG-500; Ottawa, Ontario, Canada). Agarose gel and Qubit (Life Technologies, CA, USA) were used for DNA quantity and quality assessment. DNA samples from 2 affected individuals (G2 and G4) were analyzed using a SureSelect custom NGS gene panel (MiamiOtoGenes) of 230 known and candidate genes associated with sensorineural hearing loss (Yan et al., 2016). This panel was designed to include all exons, 5’ UTRs and 3’ UTRs of the genes and covered a target size of approximately 1.598 Mbp encompassing 4422 regions. The targeted sequencing was processed at the Hussman Institute for Human Genomics (HIHG) Sequencing core, University of Miami. The SureSelectTarget Enrichment (Agilent, Santa Clara, CA, USA) of coding exons and flanking intronic sequences in-solution hybridization capture system was used according to the manufacturer’s standard protocol. Adapter sequences for the Illumina HiSeq 2000 were ligated, and the enriched DNA samples were prepared using the standard methods for the HiSeq 2000 instrument (Illumina, San Diego, CA, USA). The average insert size of 180 bp and paired-end sequencing reads were used. The sample from G3 was subjected to whole exome sequencing. The enrichment of coding exons and flanking intronic regions was performed using a solution hybrid selection method with the SureSelectXT human all exon 60 Mb v6 kit (Agilent Technologies) following the manufacturer’s standard protocol.

#### 2.2.2. Bioinformatics Analysis

Bioinformatics processing and data analysis were performed as previously reported (Yan et al., 2016). In brief, The Illumina CASAVA v1.8 pipeline was used to assemble 99 bp sequence reads. Burrows-Wheeler Aligner (BWA) was applied for alignment of sequence reads to the human reference genome (hg19) (Li & Durbin 2010), and variants were called using FreeBayes (Garrison & Marth, 2012). KGGSeq (http://pmglab.top/kggseq/) was then used for variant filtering based on quality/score read depth and minor allele frequency (MAF thresholds of 0.0005 for ADNSHL variants) as reported in dbSNP141, the National Heart, Lung, and Blood Institute Exome Sequencing Project Exome Variant Server, Seattle, WA Project (Exome Variant Server, 2012), the Genome Aggregation Database (gnomAD; https://gnomad.broadinstitute.org) and the 1000 Genome Project Database. Variants were further analyzed based on their presence and pathogenicity information in Human Gene Mutation Database (HGMD; http://www.hgmd.cf.ac.uk), the Deafness Variation Database (DVD) (deafnessvariationdatabase.org), and ClinVar (http://www.ncbi.nlm.nih.gov/clinvar/). Copy number variations (CNV) calling was performed using an R-based tool (Nord at al., 2011). This method normalizes read-depth data by sample batch and compares median read-depth ratios using a sliding-window approach. Variants meeting these criteria were further annotated according to specific standard terminology of American College of Medical Genetics and Genomics (ACMG) guidelines (Richards et al., 2015) that recommends classification of DNA variants into five categories including pathogenic, likely-pathogenic, variant of uncertain significance (VUS), likely-benign, and benign. To analyze the possible functional pathogenic effects of the missense variants, two types of prediction programs, SIFT, and Polyphen score were used. The Human Gene Mutation Database (http://www.hgmd.cf.ac.uk), the Deafness Variation Database (DVD) (deafnessvariationdatabase.org), and Clinvar (http://www.ncbi.nlm.nih.gov/clinvar/) were used as references to determine the novelty and probable pathogenicity of the allelic variations detected in our sequencing approach.

#### 2.2.3. Confirmation and Segregation Analysis

The candidate variation was confirmed via Sanger sequencing. Primer3, v. 0.4.0 (http://primer3.ut.ee) was used for primer design. Polymerase chain reaction (PCR) reactions were performed with 40–60 ng of genomic DNA and Taq DNA polymerase (Sigma) using standard protocols. PCR products were purified with Qiagen Qiaquick purification kit and bidirectionally sequenced using the ABI PRISM Big Dye Terminator Cycle Sequencing V3.1 Ready Reaction Kit and passed on the ABIPRISM 3730 DNA Analyzer (Applied Biosystems). DNA sequence analysis was performed with DNASTAR Lasergene software.

### 2.3. Electrophysiological Evaluation Procedures

All eCAP measures were conducted using the Advanced Neural Response Telemetry function via the Custom Sound EP (v. 5.2) software interface (Cochlear Ltd, Sydney, NSW, Australia). For measuring the eCAP AGF and RRF, the stimulus was a symmetric, cathodic-leading, biphasic pulse with a pulse phase duration of 25 μs/phase. The presentation rate of the stimulus was 15 Hz. For measuring *τ*_3_, the probe was the single pulse as that described for measuring the eCAP AGF and RRF. The masker stimulus was a train of biphasic pulses with the same characteristics as those of the single-pulse stimulus. The duration of this masker pulse train was 100 ms with a carrier rate of 900 pulses per second (pps) per channel. The presentation rate of the masker pulse train and the probe pulse was 2 Hz. Other recording parameters included an amplifier gain of 50 dB, a sampling delay of 122 μs, an effective sampling rate of 20 kHz, and 50 sweeps per averaged eCAP response. The stimulus was presented to individual CI electrodes in a monopolar-coupled configuration via a N6 sound processor that was connected to a programming pod.

For eCAP AGF and RRF measures, electrode locations tested in children with NSCNs and postlingually deafened adult CI users were the same as those tested in G1 and G4. In children with CND, three electrodes across electrode array with measurable eCAPs were tested. In this study, these electrodes were considered as the “basal”, the “middle,” and the “apical” electrode based on their relative locations among electrodes with measurable eCAPs. Details for selecting these electrode locations in children with CND were reported in He et al. (2020). For *τ*_3_ estimation, electrode locations tested in G1 and G4 were similar with those tested in other postlingually deafened adult CI users with various etiologies for hearing loss.

#### 2.3.1. Slope of the Amplitude Growth Function

Detailed approaches used to measure the eCAP AGF and estimate slope have been reported in Skidmore et al. (2021). Briefly, the eCAP AGF was obtained using the forward-masking-paradigm. The probe pulse was initially presented at 10 clinical unit levels (CLs) lower than the maximum comfortable level (C level) and decreased by 1 CL for at least five consecutive measurements. Subsequently, the stimulation level of the probe pulse systematically decreased in steps of 5 CLs until no eCAP response could be visually identified. The stimulation level was then increased in steps of 1 CL until at least five consecutive eCAPs were measured using this small step size. In this test, the masker pulse was always presented at 10 CLs higher than the probe pulse. The eCAP AGF was obtained by plotting the eCAP amplitude as a function of stimulation level. The slope of the eCAP AGF was estimated using the “Window method” (Skidmore et al., 2022) at each electrode location listed in Table 2.

#### 2.3.2. Absolute Refractory Period

Detailed approaches used to measure the eCAP RRF and estimate *t*_*0*_ have been reported in our previous studies (He et al., 2018; Skidmore et al., 2021). Briefly, the eCAP RRF was obtained using a modified template subtraction method. A series of eCAPs were recorded as the masker-probe-interval (MPI) was systematically increased from 0.40 to 10.0 ms. The eCAP RRF was obtained by plotting normalized eCAP amplitudes (re: the eCAP amplitude measured at the MPI of 10 ms) as a function of MPI. An estimate of *t*_*0*_ was found using statistical modeling with an exponential decay function. For each test ear, *t*_*0*_ s measured at three electrode locations that are listed in Table 2.

#### 2.3.3. The Speed of Neural Adaptation Recovery

The recovery speed from neural adaptation of the CN fibers (i.e., *τ*_3_) was measured using the same method as that reported in He et al. (2022). Briefly, eCAPs evoked by the probe pulse presented at different time intervals after the cessation of the last pulse of the masker pulse train were measured. Time intervals tested in this study included 1.054, 2, 4, 8, 16, 32, 64, 128 and 256 ms. To obtain the adaptation recovery function, normalized eCAP amplitudes (re: the eCAP amplitude measured at the MPI of 256 ms) were plotted as a function of tested time interval. *τ*_3_ was estimated based on the adaptation recovery function using a mathematical model with up to three exponential components. Details of this mathematical model have been reported in He et al. (2022).

## 3. Results

### 3.1 Clinical Features of the American Family

The family spanning five generations showed a typical autosomal dominant inheritance pattern of nonsyndromic hearing loss (Figure 1). There was no vestibular dysfunction or other clinical abnormalities indicating syndromic hearing loss. Audiological assessments of the patients revealed bilateral, sensorineural hearing loss that began in the early or mid-30s affecting all genders that progressed to profound hearing loss by age 50-60 involving all frequencies.

**Figure 1.**
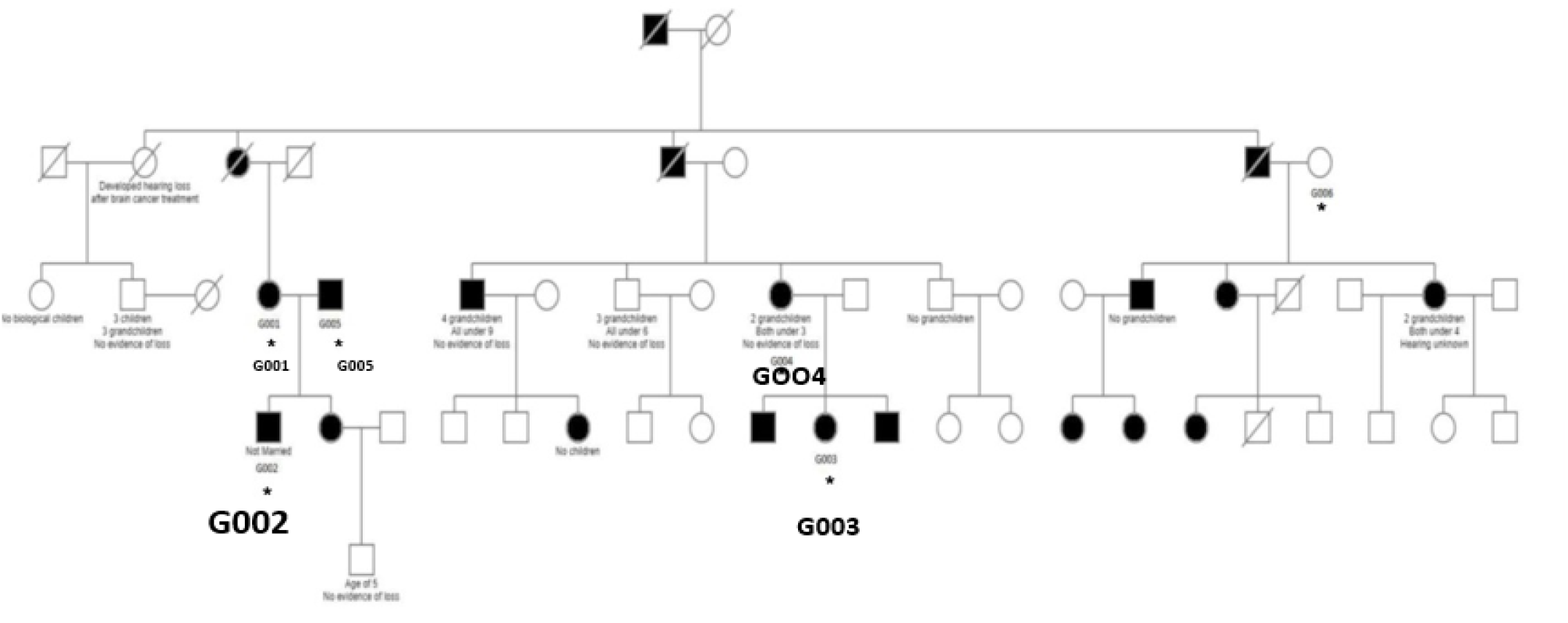
Pedigree of the family with ADSNHL. The pedigree of the five-generation family with the Gln246Leu variant in the *ACTG1* gene. Filled symbols represent the affected individuals.

### 3.2. Genetic Results

#### 3.2.1. Mutations Detection

In this study, we ascertained a five-generation family with progressive ADNSHL. We used our targeted custom MiamiOtoGenes panel containing 230 hearing loss-associated genes to analyze DNA samples of 2 (G2 and G4) affected family members and sample of 1 patient (G3) was subjected to whole exome sequencing (WES). We then used KGGSeq (http://pmglab.top/kggseq/) for variant filtering based on quality/score read depth according to the autosomal dominant inheritance model, functional significance (i.e., nonsense, missense, or splice site), single-nucleotide variants shared by the 3 affected individuals and minor allele frequency (MAF) thresholds of 0.0005 for ADNSHL variants as reported in public databases. Only 1 heterozygous variant shared by the 3 patients analyzed by NGS was identified using this simple filtering strategy. This sequence change is located in the *ACTG1* (hg19) gene on chromosome 17q25.3 at 7,9478,279, c.737A>T (NM_001614.5) leading to (p.Gln246Leu) (P63261). The identified variant (rs1568061110) has not been published as a pathogenic variant, nor has it been reported as a benign variant to our knowledge. The variant is not observed in large population cohorts (gnomAD). The results of Sanger sequencing analysis revealed that all of the available affected family members were heterozygous for this variant, while it was not observed in family members with normal hearing (Fig 2A, B). The residue glutamine at the position 246 is highly conserved across the species (Fig 2C) and the evolutionary conservation of the involved amino acid is further corroborated by genomic evolutionary rate profiling conservation score (GERP) and PhastCons analysis. Moreover, the observed Gln to Leu amino acid change at position 246 is predicted to be damaging according to PolyPhen, MutationTaster, MutationAssessor, LRT, FATHMM, GERP+, VEST3, PROVEAN, MetaSVM_pred, MetaLR_pred and phastCons. *In silico* deleterious prediction analysis indicated that the identified variant was potentially deleterious. The characteristics are summarized in Table 3.

**Table 3.** Scores and predicted effects of the observed Gln to Leu amino acid change at position 246 determined using different tools.

| Tool | Score | Prediction |
| --- | --- | --- |
| Polyphen2 HDIV | 0.646000 | probably damaging |
| Polyphen2 HVAR | 0.915000 | probably damaging |
| LRT | 0.000002 | deleterious |
| MutationTaster | 0.588140 | disease_causing |
| MutationAssessor | 3.895000 | low |
| FATHMM | 3.470000 | disease_causing |
| VEST3 | 0.742000 |  |
| PROVEAN | -4.710000 | disease_causing |
| MetaSVM_pred |  | disease_causing |
| MetaLR_pred |  | disease_causing |
| GERP++_NR | 4.560000 |  |
| phastCons7way_vertebrate | 0.993000 |  |

**Figure 2.**
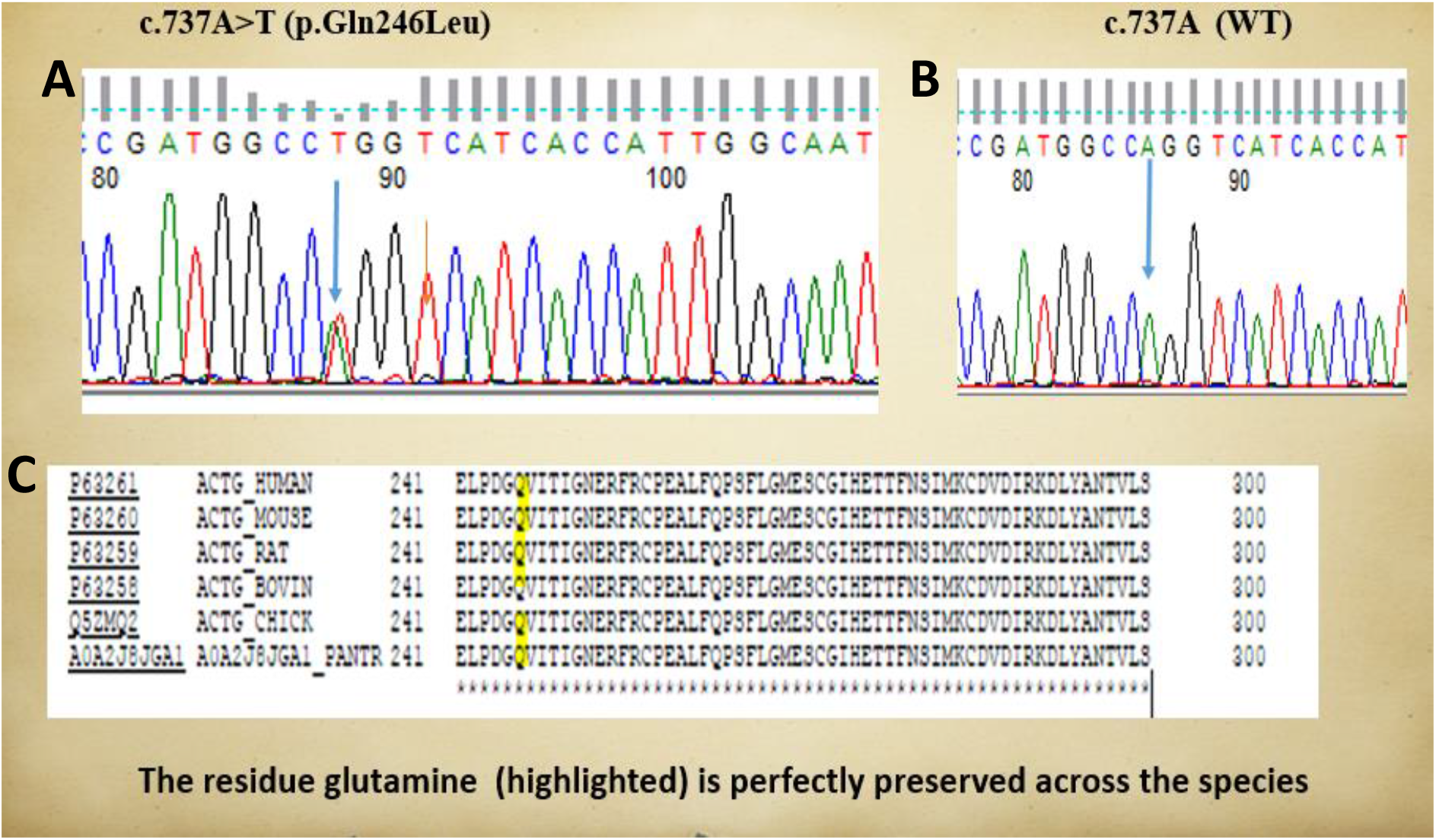
DNA sequencing chromatograms. DNA sequencing chromatograms show the c.737A>T (p.Gln246Leu) variant in *ACTG1* in a heterozygous state (A), while normal hearing member has wild-type allele (B). The residue Q (highlighted) is perfectly preserved across the species

#### 3.2.2. Structural Modeling of Gln246Leu

The human cytoplasmic actomyosin complex based on the cryo-EM structure (pdb id: 5jlh), shows γ-Actin chains wrapped by Tropomyosin α-3 helices. The Gln246 residue in VDW representation (Figure 3A) and other neighboring important residues, including the variant Gln246Leu, at the interface between two γ-Actin chains are shown as sticks (Figure 3B). We have performed the variant modeling with Chimera (Pettersen et al., 2004) and the structure was rendered with VMD (Humphrey et al., 1996).

**Figure 3.**
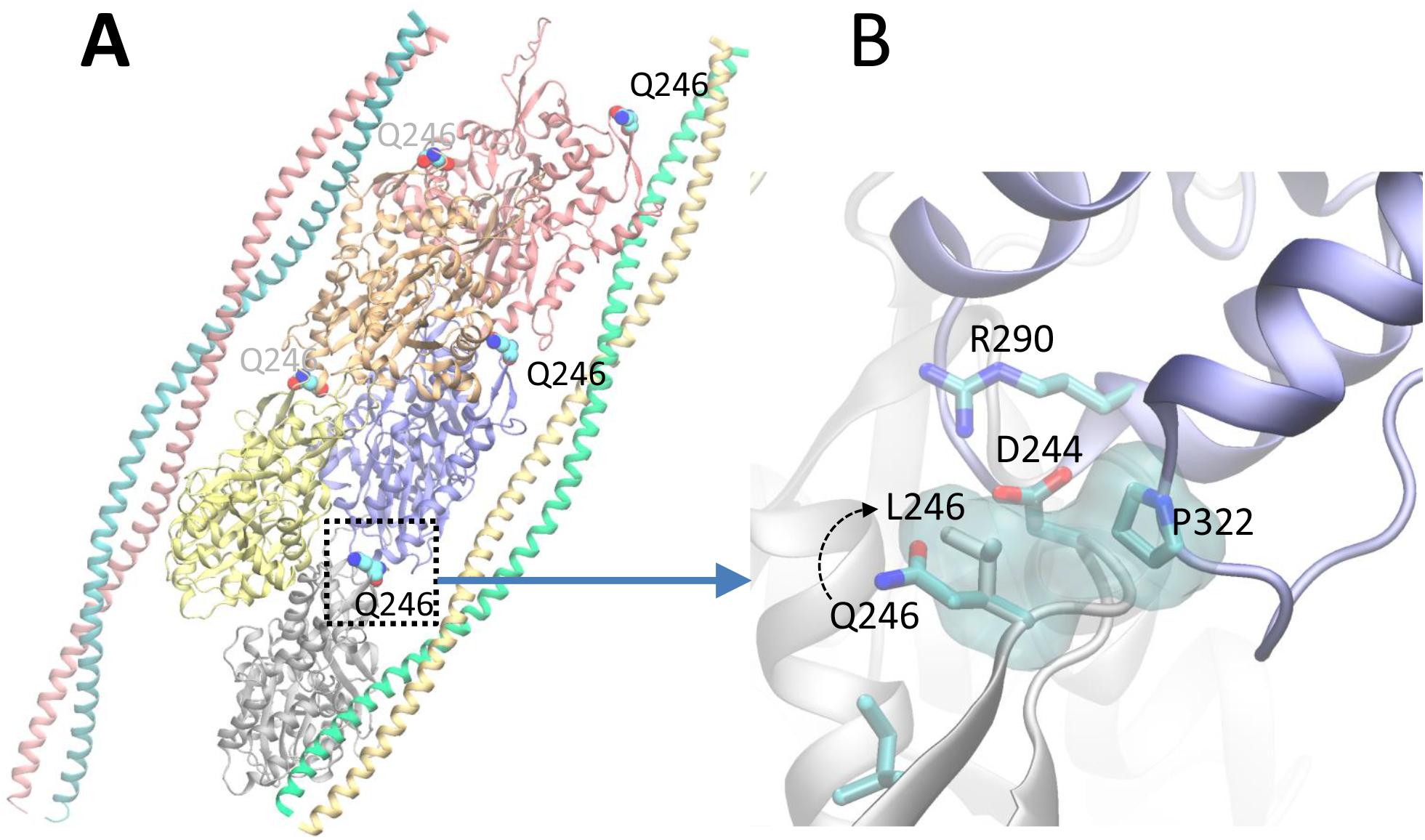
Human cytoplasmic actomyosin complex based on the cryo-EM structure (pdb id: 5jlh), showing γ-Actin chains wrapped by Tropomyosin α-3 helices. The Gln246 residue shown in VDW representation (A); neighboring residues including the Gln246Leu, at the interface between two γ-Actin chains are shown as sticks (B). The variant was modeled with Chimera (Pettersen et al., 2004) and the structure was rendered with VMD (Humphrey et al., 1996).

In the cytoplasmic actomyosin complex, the residue Gln246 lies at the interface between γ-Actin chains. As shown in Fig 3B, Asp244 in one chain makes an ionic interaction with Arg290 of the other chain. The sidechain of Gln246 is in the proximity of Asp244 in the wild type. The Gln246Leu variant creates a non-specific hydrophobic contact between Leu246 and Pro322, and possibly affects proper orientation of Asp244. Therefore, the Gln246Leu variant causes significant changes at the interfacial interactions between two γ-Actin chains.

### 3.3. Electrophysiological Measurement Results

For each eCAP measure, results measured at different electrode locations in G1 and G4 are listed in Tables 4 and 5. In addition, their results were compared with 24 implanted children with CND, 32 implanted children with idiopathic sensorineural hearing loss and NSCNs, and 43 postlingually deafened adult CI users with various etiologies of hearing loss. Detailed demographic information, including individual patients’ etiologies of hearing loss, and study results of these three comparative implanted patient populations have been reported in our previous studies (He et al., 2018; Skidmore et al., 2021; He et al., 2022), and were not re-stated here.

**Table 4.**
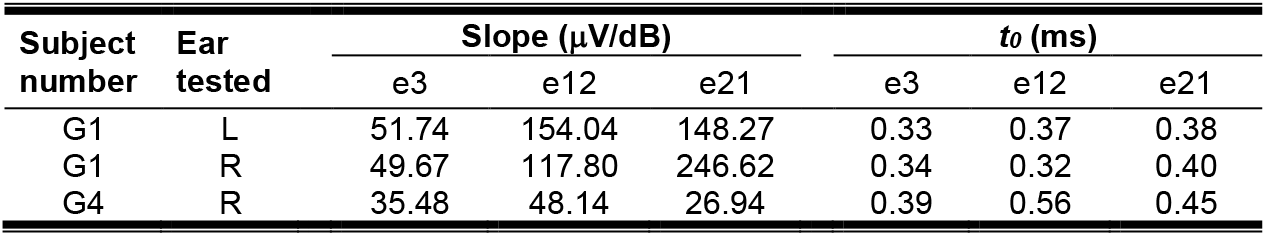
Slopes of the electrically evoked compound action potential amplitude growth function and absolute refractory periods (i.e., *t*_0_*s*) measured in G1 and G4. L: left ear, R: right ear, e3: electrode 3, e12: electrode 12, e21: electrode 21.

**Table 5.** *τ*_3_s measured in G1 and G4 at four electrode locations. L: left ear. R: right ear, e3: electrode 3, e9: electrode 9, e12: electrode 12, e15: electrode 15, e21: electrode 21.

| Subject<br>number | Ear tested | $\tau_3$ (ms) | | | |
| --- | --- | --- | --- | --- | --- |
|  |  | e3 | e9/e12 | e15 | e21 |
| G1 | L | 49.08 | 6.11 | 6.51 | 5.00 |
| G1 | R | 29.55 | 7.06 | 20.00 | 6.47 |
| G4 | R | 13.74 | 20.74 | 60.00 | 20.61 |

#### 3.3.1. Slope of the eCAP Amplitude Growth Function

Data listed in Table 4 clearly show that slopes of the eCAP AGF measured in G1 and G4 varied substantially and demonstrated different trends. Specifically, slopes measured in both ears of G1 increased as the stimulating electrode moved from the base (i.e., electrode 3) to more apical locations of the cochlea (i.e., electrode 12 and electrode 21). This trend was not observed in results measured in G4. In addition, slopes measured in both ears of G1 were substantially larger than those measured in G4 at all three electrode locations.

The top row of Figure 5 depicts results measured in these two participants and the means and standard deviations of slopes measured in three comparative CI patient populations. It is apparent that slopes measured at the middle-array and the apical electrodes in both ears of G1 were substantially larger than those measured in all three CI patient populations. In contrast, slopes measured in G4 were smaller than the averaged slopes measured in implanted children with NSCNs (86.23-83.19 μV/dB) and other adult CI users (42.18-60.56 μV/dB) and appeared to be similar with those measured in implanted children with CND (43.23-50.00 μV/dB).

**Figure 4.**
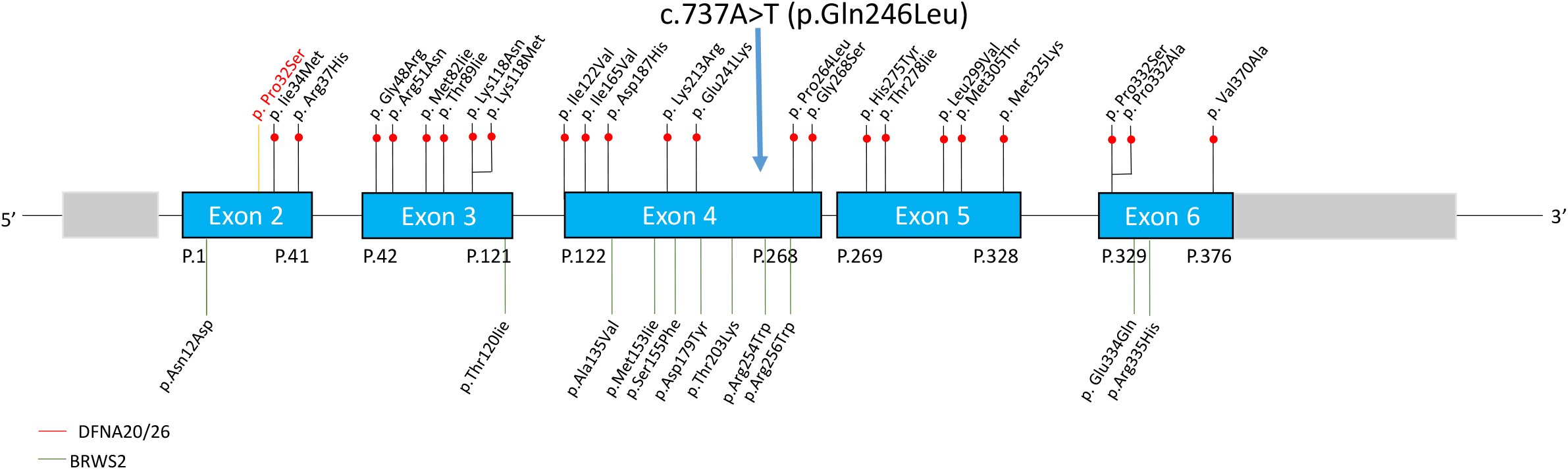
Schematic representation of the full length *ACTG1* gene showing the six exons. The mutations associated with DFNA20/26 are indicated above the gene and those linked to Baraitser –Winter syndrome are represented below the exons

**Figure 5.**
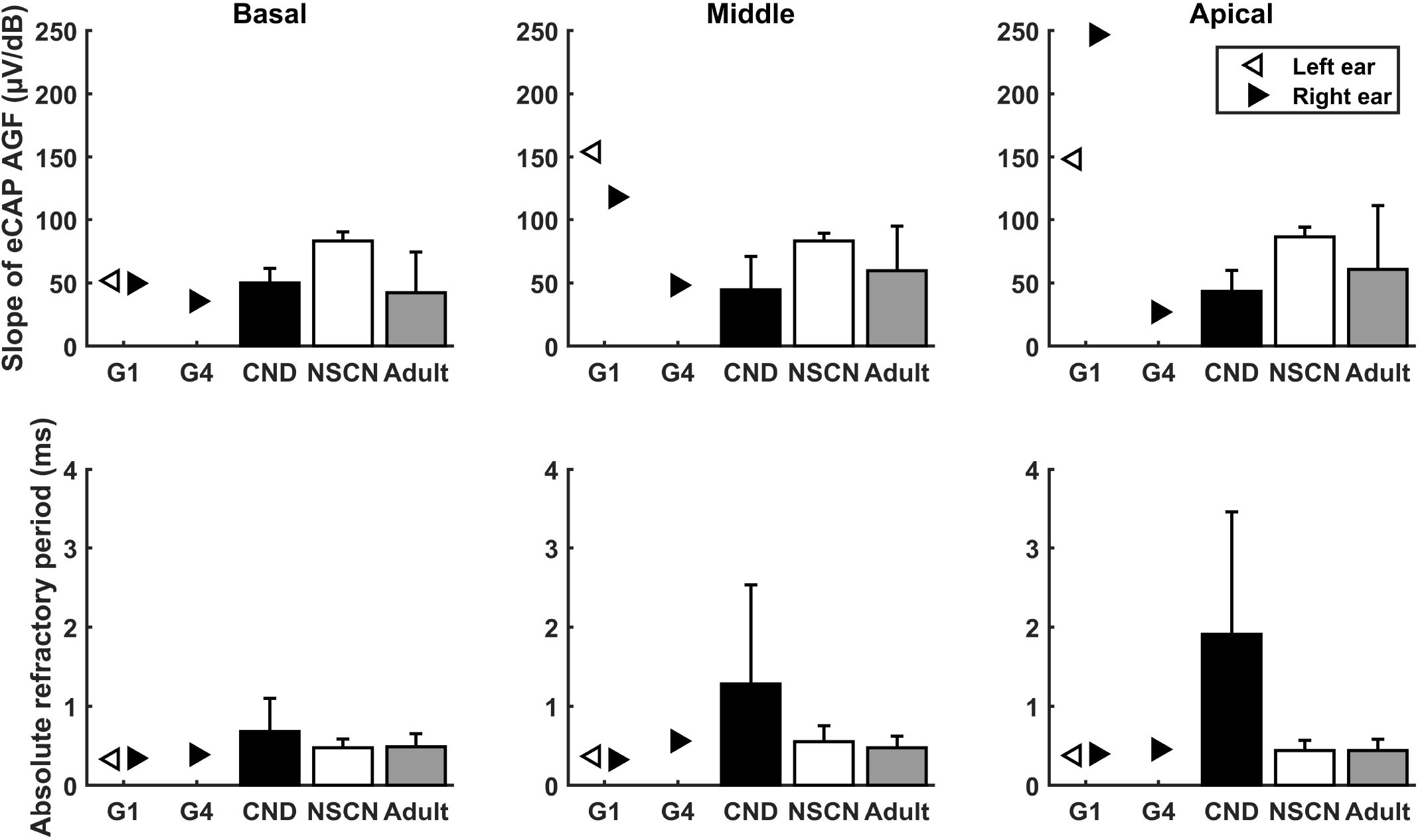
Results of slope of the eCAP AGF (top row) and absolute refractory period of the eCAP RRF (bottom row) measured in G1 and G4, along with the means and standard deviations of the results from the three comparative CI patient populations.

#### 3.3.2. Absolute Refractory Period

Compared with slopes of the eCAP AGF, *t*_*0*_s measured these two participants (Table 4) show much less variations between participants. In addition, unlike slopes of the eCAP AGF, *t*_*0*_s measured in these two participants do not show any apparent trend that was associated with electrode location.

The bottom row of Figure 5 shows *t*_*0*_s measured in these two participants and the means and the standard deviations of results measured in three CI patient populations. These data show that children with CND clearly had much longer *t*_*0*_s than other CI users. *t*_*0*_s measured in all electrode locations in both ears of G1 were shorter than the means of *t*_*0*_s measure in implanted children with CND (0.68-1.91 ms), implanted children with NSCNs (0.44-0.55 ms), and other adult CI users (0.44-0.49 ms). In contrast, *t*_*0*_s measured in G4 were similar with those measured in implanted children with NSCNs and other adult CI users but smaller than those measured in implanted children with CND.

#### 3.3.3. The speed of Neural Adaptation Recovery

*τ*_3_ s measured at different electrode locations in G1 and G4 are listed in Table 5. Similar to slopes of the eCAP AGF, substantial variations in *τ*_3_ were observed for results measured at different electrodes within the same participants as well as between these two participants. In both ears of G1, *τ*_3_ measured at the basal electrode location was substantially longer than those measured at all other electrode locations. This data trend was not observed for results measured in G4.

Figure 6 shows the comparison between results measured in these two participants and those estimated for other adult CI user with different etiologies. *τ*_3_s measured at all except for the basal electrode location were much shorter than the mean of *τ*_3_s (48.72-52.74 ms) measured in other postlingually deafened adult CI users. *τ*_3_ s measured in G4 are within the same range as those measured in other adult CI users.

**Figure 6.**
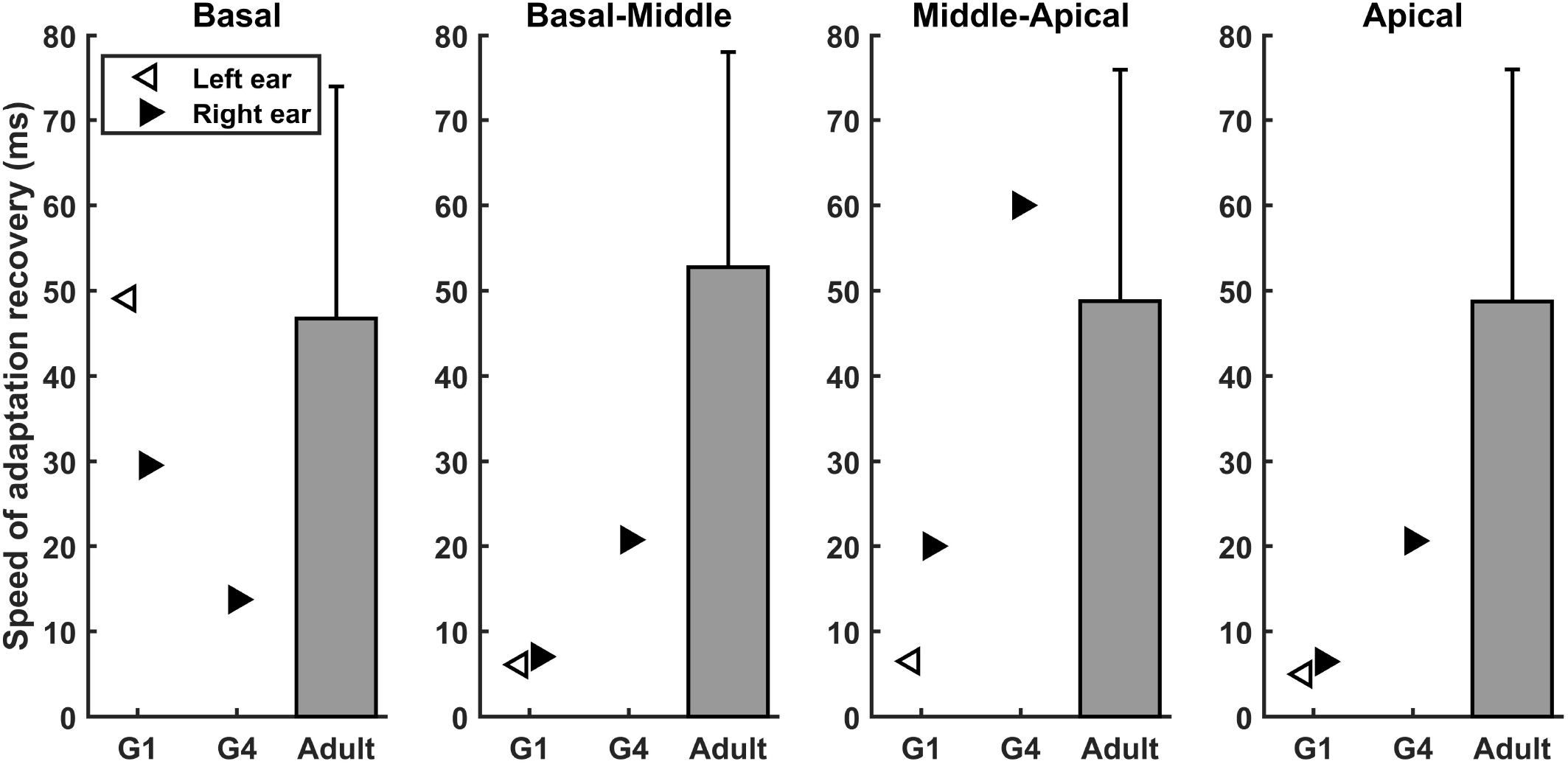
Results of speeds of adaptation recovery measured inG1 and G4, along with the means and standard deviations of the data from the comparative adult cochlear implant user group.

## 4. Discussion

The present study extended the clinical features of hearing loss patients with *ACTG1* variants by identification and characterization of the missense Gln246Leu variant in *ACTG1*. Furthermore, this preliminary study incorporated electrophysiological measures of the eCAP to explore the direct link between this genetic variant and the responsiveness of the CN to electrical stimulation in human listeners. Our limited results showed substantial variations in different eCAP measures in two participants with this missense change. Overall, these limited results highlight the importance of evaluating the ability of the peripheral auditory system to respond to electrical stimulation in individual CI users with different genetic profiles.

### Variants reported in the *ACTG1* gene

Up to now, more than 30 mutations in the *ACTG1* gene have been reported (Figure 4). Several mutations in the *ACTG1* gene were found to cause ADNSHL linked to the DFNA20/26 locus (Rendtorff et al., 2006; Van Wijk et al., 2003). For example, Van Wijk et al. (2003) identified the c.833C>T (p.Thr278Ile) mutation in the gene encoding cytoplasmic ɤ-1-actin (*ACTG1*) in a Dutch family with ADNSHL (DFNA20/26 locus). Rendtorff et al. (2006) also found a missense mutation (c.1109T>C; p.Val370Ala) in *ACTG1* in a Norwegian DFNA20/26 family.

So far, only missense mutations have been identified and located in various binding domain. Most of the *ACTG1* mutations are “private”, with the exception of three DFNA20/26 associated mutations (K118M, E241K, and L299V) that were identified in at least two families. The K118M mutation in *ACTG1* has been reported in four families and is thus the most common mutation. In addition, a mutation involving the same residue (K118N) has also been identified in a Spanish family. So far, at least five of the mutations identified in DFNA20/26 families worldwide have been found to carry mutation involving the residue K118, suggesting that codon K118 of *ACTG1* may represent a mutational hot spot (Wang et al., 2018) that can be used as the key screening site of the ACTG1 gene detection and the target of gene therapy in the future.

### Responsiveness of the CN to Electrical Stimulation

The present work explored the functional, phenotypic effect of this *ACTG1* missense variant on the responsiveness of the CN to electrical stimulation. Decline in CN responsiveness in implanted patients with *ACTG1* missense variant was not expected because the primary targeting site of this nucleotide change is believed to be the sensory partition (organ of Corti and synapse) of the cochlea. Nevertheless, eCAP results measured in the two participant who carry the same missense variant showed substantial variations and an inconsistency in alignment with expected results. Specifically, slopes of the eCAP AGF measured in both ears of G1 and in other adult CI users gradually increased as the stimulating electrode moved toward more apical locations within the cochlea (Figure 5). This increase can be due to the gradually reduced diameter of the cochlear turn toward the apical end, the distance between the electrode and the local CN fibers is smaller for more apical electrode locations than for electrodes near basal regions. In addition, it has been shown that neural structures at the apex tend to have better neural survival or be healthier than those located at more basal region of the cochlea in typical listeners with sensorineural hearing loss (Zimmermann et al., 1995). Both factors can lead larger slopes measured at more apical electrode locations (e.g., Zimmermann et al., 1995). More importantly, slopes measured at the middle-array and the apical electrode location were substantially larger than any other CI patient populations reported in this study (Figure 5). Results of *τ*_3_ measured in both ear of G1 were consistent with the slope data in terms of the expected CN responsiveness. Finally, *t*_*0*_s measured in all electrode locations in both ears of G1 were shorter than results measured in all three comparative CI patient populations. Overall, these results suggest robust responsiveness of the electrically stimulated CN in G1, which aligns with the expected results.

In contrast, the electrode-location associated data trend in slope or *τ*_3_ was not observed in the results measured in the right ear of G4 (Figure 5). As a matter of fact, slopes measured in G4 appeared to be similar with those measured in implanted children with CND, a patient population with damages in the CN. *t*_*0*_s and *τ*_3_s measured in G4 were similar with those measured in implanted children with NSCNs and/or other adult CI users. Overall, these results failed to support the idea that this missense variant does not affect responsiveness of the CN to electrical stimulation in human CI users.

Due to the extremely limited demographic and audiologic data, factors leading to these inconsistent results remain unknown. We can only speculate that the difference in duration of deafness might have, at least partially, contributed to the inconsistency in eCAP results measured in G1 and G4. Specifically, the self-reported duration of deafness of G4 was 56.5 years, which is much longer than that reported by G1 (i.e., 30.7 years). In animal models, longer duration of deafness have been shown to lead to greater functional (e.g., altered spontaneous activity, increased absolute refractory period) and anatomical deterioration (demyelination, loss of peripheral axons and SGNs) of the CN (e.g., Shepherd et al., 2004). These deteriorations will reduce the CN responsiveness to electrical stimulation as measured using the eCAP. In a separate project, the annual noise exposure was assessed and quantified using Noise Exposure Questionnaire (Johnson et al., 2017). The scores for G1 and G4 were 77 and 72, respectively. Therefore, the difference in eCAP results between G1 and G4 is unlikely due to the difference in the amount of annual noise exposure.

Even though the inconsistency in eCAP results measured in G1 and G4 is disappointing and somewhat confusing at this point, these results do suggest the importance of 1) evaluating auditory function for individual patients even with the same genetic profiles, and 2) incorporating patient-specific factors when interpreting and utilizing genetic results for patient care.

## 5. Conclusion

The missense c.737A>T (p. Gln246Leu) variant in *ACTG1* (NM_001614.5) co-segregated with hearing loss and the most likely to be the genetic cause of ADNSHL in a five-generation American family. Different functional, phenotypic results can be observed in patients with the same genetic variant, which suggests the importance of assessing the functional status of the CN for individual CI users. Further studies with more participants are warranted to test the feasibility of accurately predicting the responsiveness of the electrically stimulated CN based on genetic profiles and patient-specific factors in human CI users.

## Data Availability

Please contact the corresponding authors to discuss access to the data presented in this study.

## Acknowledgements

This work was supported by the R01 grants (R01 DC005575 and R01 DC012115) from the National Institutes of Health/National Institute on Deafness and Other Communication Disorders awarded to Xue Zhong Liu, as well as the R01 (R01DC017846) from the National Institute on Deafness and Other Communication Disorders and the R01 grant (R01DC016038) from the National Institute on Deafness and Other Communication Disorders and the National Institute of General Medical Sciences awarded to Shuman He.

## Data Availability Statement

The data that support the findings of this study are available on request from the corresponding authors, XHL and SH.

## References

DeRosier, D. J., & Tilney, L. G. (1989). The structure of the cuticular plate, an in vivo actin gel. Journal of Cell Biology, 109(6), 2853–2867. https://doi.org/10.1083/jcb.109.6.2853

Elfenbein, J. L., Fisher, R. A., Wei, S., Morell, R. J., Stewart, C., Friedman, T. B., & Friderici, K. (2001). Audiologic aspects of the search for DFNA20: A gene causing late-onset, progressive, sensorineural hearing loss. Ear and Hearing, 22(4), 279–288. https://doi.org/10.1097/00003446-200108000-00003

Ferron, F., Rebowski, G., Lee, S. H., & Dominguez, R. (2007). Structural basis for the recruitment of profilin–actin complexes during filament elongation by Ena/Vasp. The EMBO Journal, 26(21), 4597–4606. https://doi.org/10.1038/sj.emboj.7601874

Garrison, E., & Marth, G. (2012). Haplotype-based variant detection from short-read sequencing. https://arxiv.org/abs/1207.3907

He, S., Shahsavarani, B. S., McFayden, T. C., Wang, H., Gill, K. E., Xu, L., Chao, X., Luo, J., Wang, R., & He, N. (2018). Responsiveness of the electrically stimulated cochlear nerve in children with cochlear nerve deficiency. Ear & Hearing, 39(2), 238–250. https://doi.org/10.1097/aud.0000000000000467

He, S., Xu, L., Skidmore, J., Chao, X., Jeng, F. C., Wang, R., Luo, J., & Wang, H. (2020). The effect of Interphase Gap on neural response of the electrically stimulated cochlear nerve in children with cochlear nerve deficiency and children with normal-sized cochlear nerves. Ear and Hearing, 41(4), 918–934. https://doi.org/10.1097/aud.0000000000000815

He, S., Skidmore, J., Carter, B. L., Lemeshow, S., & Sun, S (2022). Postlingually Deafened Adult Cochlear Implant Users With Prolonged Recovery From Neural Adaptation at the Level of the Auditory Nerve Tend to Have Poorer Speech Perception Performance. Ear and Hearing, 10–1097.

Hilgert, N., Smith, R., & Camp, G. (2009). Function and expression pattern of non-syndromic deafness genes. Current Molecular Medicine, 9(5), 546–564. https://doi.org/10.2174/156652409788488775

Hirokawa, N., & Tilney, L. G. (1982). Interactions between actin filaments and between actin filaments and membranes in quick-frozen and deeply etched hair cells of the chick ear. Journal of Cell Biology, 95(1), 249–261. https://doi.org/10.1083/jcb.95.1.249

Humphrey, W., Dalke, A., & Schulten, K. (1996). VMD: Visual molecular dynamics. Journal of Molecular Graphics, 14(1), 33–38. https://doi.org/10.1016/0263-7855(96)00018-5

Johnson, T. A., Cooper, S., Stamper, G. C., & Chertoff, M. (2017). Noise exposure questionnaire: A tool for quantifying annual noise exposure. Journal of the American Academy of Audiology, 28(01), 14–35. https://doi.org/10.3766/jaaa.15070

Kovar, D. R., Harris, E. S., Mahaffy, R., Higgs, H. N., & Pollard, T. D. (2006). Control of the Assembly of ATP-and ADP-actin by Formins and Profilin. Cell, 124(2), 423–435. https://doi.org/10.1016/j.cell.2005.11.038

Li, H., & Durbin, R. (2009). Fast and accurate short read alignment with Burrows-wheeler transform. Bioinformatics, 25(14), 1754–1760. https://doi.org/10.1093/bioinformatics/btp324

Long, C. J., Holden, T. A., McClelland, G. H., Parkinson, W. S., Shelton, C., Kelsall, D. C., & Smith, Z. M. (2014). Examining the electro-neural interface of cochlear implant users using psychophysics, CT scans, and speech understanding. Journal of the Association for Research in Otolaryngology, 15(2), 293–304. https://doi.org/10.1007/s10162-013-0437-5

Michelot, A., & Drubin, D. G. (2011). Building distinct actin filament networks in a common cytoplasm. Current Biology, 21(14), R560–R569. https://doi.org/10.1016/j.cub.2011.06.019

Morell, R. J., Friderici, K. H., Wei, S., Elfenbein, J. L., Friedman, T. B., & Fisher, R. A. (2000). A new locus for late-onset, progressive, hereditary hearing loss DFNA20 maps to 17q25. Genomics, 63(1), 1–6. https://doi.org/10.1006/geno.1999.6058

Nord, A. S., Lee, M., King, M.-C., & Walsh, T. (2011). Accurate and exact CNV identification from targeted high-throughput sequence data. BMC Genomics, 12(1). https://doi.org/10.1186/1471-2164-12-184

Pennings, R., Huygen, P., Van Camp, G., & Cremers, C. (2003). A review of progressive phenotypes in non-syndromic autosomal dominant hearing impairment. Audiological Medicine, 1(1), 47–55. https://doi.org/10.1080/16513860310003085

Pettersen, E. F., Goddard, T. D., Huang, C. C., Couch, G. S., Greenblatt, D. M., Meng, E. C., & Ferrin, T. E. (2004). UCSF chimera -- a visualization system for exploratory research and analysis. Journal of Computational Chemistry, 25(13), 1605–1612. https://doi.org/10.1002/jcc.20084

Pollard, T. D., & Cooper, J. A. (2009). Actin, a central player in cell shape and Movement. Science, 326(5957), 1208–1212. https://doi.org/10.1126/science.1175862

Ramekers, D., Versnel, H., Strahl, S. B., Klis, S. F., & Grolman, W. (2015). Temporary neurotrophin treatment prevents deafness-induced auditory nerve degeneration and preserves function. Journal of Neuroscience, 35(36), 12331–12345. https://doi.org/10.1523/jneurosci.0096-15.2015

Rendtorff, N. D., Zhu, M., Fagerheim, T., Antal, T. L., Jones, M. P., Teslovich, T. M., Gillanders, E. M., Barmada, M., Teig, E., Trent, J. M., Friderici, K. H., Stephan, D. A., & Tranebjærg, L. (2006). A novel missense mutation in ACTG1 causes dominant deafness in a Norwegian DFNA20/26 family, but ACTG1 mutations are not frequent among families with hereditary hearing impairment. European Journal of Human Genetics, 14(10), 1097–1105. https://doi.org/10.1038/sj.ejhg.5201670

Richards, S., Aziz, N., Bale, S., Bick, D., Das, S., Gastier-Foster, J., Grody, W. W., Hegde, M., Lyon, E., Spector, E., Voelkerding, K., & Rehm, H. L. (2015). Standards and guidelines for the interpretation of sequence variants: A joint consensus recommendation of the American College of Medical Genetics and genomics and the Association for Molecular Pathology. Genetics in Medicine, 17(5), 405–424. https://doi.org/10.1038/gim.2015.30

Rivière J.B., van Bon B.W., Hoischen A., Kholmanskikh S.S., O’Roak B.J., Gilissen C., Gijsen S., Sullivan C.T., Christian S.L., Abdul-Rahman O.A., Atkin J.F., Chassaing N., Drouin-Garraud V., Fry A.E., Fryns J.P., Gripp K.W., Kempers M., Kleefstra T., Mancini G.M., Nowaczyk M.J., van Ravenswaaij-Arts C.M., Roscioli T., Marble M., Rosenfeld J.A., Siu V.M., de Vries B.B., Shendure J., Verloes A., Veltman J.A., Brunner H.G., Ross M.E., Pilz D.T., Dobyns W.B. (2012). De novo mutations in the actin genes ACTB and ACTG1 cause Baraitser-Winter syndrome. Nature genetics, 44(4), 440–444.

Shearer, A. E., Hildebrand, M. S., and Smith, R. J. (1993). “Hereditary hearing loss and deafness overview,” in GeneReviews®, eds M. P. Adam, H. H. Ardinger, R. A. Pagon, S. E. Wallace, L. J. H. Bean, K. Stephens, et al. (Seattle, WA: University of Washington).

Shepherd, R. K., Roberts, L. A., & Paolini, A. G. (2004). Long-term sensorineural hearing loss induces functional changes in the rat auditory nerve. European Journal of Neuroscience, 20(11), 3131–3140. https://doi.org/10.1111/j.1460-9568.2004.03809.x

Sheterline, P., Clayton, J., & Sparrow, J. C. (1998). Actin. Oxford University Press.

Skidmore, J., Ramekers, D., Colesa, D. J., Schvartz-Leyzac, K. C., Pfingst, B. E., & He, S. (2022). A broadly applicable method for characterizing the slope of the electrically evoked compound action potential amplitude growth function. Ear and Hearing, 43(1), 150–164. https://doi.org/10.1097/aud.0000000000001084

Skidmore, J., Xu, L., Chao, X., Riggs, W. J., Pellittieri, A., Vaughan, C., Ning, X., Wang, R., Luo, J., & He, S. (2021). Prediction of the functional status of the cochlear nerve in individual cochlear implant users using machine learning and electrophysiological measures. Ear and Hearing, 42(1), 180–192. https://doi.org/10.1097/aud.0000000000000916

Teig, E. (1968). Hereditary progressive perceptive deafness in a family of 72 patients. Acta Oto-Laryngologica, 65(1-6), 365–372. https://doi.org/10.3109/00016486809120977

Tilney, L. G., Egelman, E. H., DeRosier, D. J., & Saunder, J. C. (1983). Actin filaments, stereocilia, and hair cells of the bird cochlea. II. packing of actin filaments in the stereocilia and in the cuticular plate and what happens to the organization when the stereocilia are bent. Journal of Cell Biology, 96(3), 822–834. https://doi.org/10.1083/jcb.96.3.822

Van Wijk, E., Krieger, E., & Kemperman, M. H. (2003). A mutation in the gamma actin 1 (ACTG1) gene causes autosomal dominant hearing loss (DFNA20/26). Journal of Medical Genetics, 40(12), 879–884. https://doi.org/10.1136/jmg.40.12.879

Wang, L., Yan, D., Qin, L., Li, T., Liu, H., Li, W., Mittal, R., Yong, F., Chapagain, P., Liao, S., & Liu, X. (2018). Amino acid 118 in the deafness causing (DFNA20/26) ACTG1 gene is a mutational hot spot. Gene Reports, 11, 264–269. https://doi.org/10.1016/j.genrep.2018.04.011

Yan D., Tekin D., Bademci G., Foster J. 2nd, Cengiz F.B., Kannan-Sundhari A., Guo S., Mittal R., Zou B., Grati M., Kabahuma R.I., Kameswaran M., Lasisi T.J., Adedeji W.A., Lasisi A.O., Menendez I., Herrera M., Carranza C., Maroofian R., Crosby A.H., Bensaid M., Masmoudi S., Behnam M., Mojarrad M., Feng Y., Duman D., Mawla A.M., Nord A.S., Blanton S.H., Liu X.Z., Tekin M.(2016). Spectrum of DNA variants for nonsyndromic deafness in a large cohort from multiple continents. Hum Genet, 135(8):953–61. doi: 10.1007/s00439-016-1697-z.

Yang, T., & Smith, R. (2000). A novel locus DFNA 26 maps to chromosome 17q25 in two unrelated families with progressive autosomal dominant hearing loss. American journal of human genetics, 67(4), 300–300.

Zimmermann, C. E., Burgess, B. J., & Nadol, J. B. (1995). Patterns of degeneration in the human cochlear nerve. Hearing Research, 90, 192–201. https://doi.org/10.1016/0378-5955(95)00165-1

